# Sex-stratified RNA-seq analysis reveals traumatic brain injury-induced transcriptional changes in the female hippocampus conducive to dementia

**DOI:** 10.1101/2022.08.24.22279173

**Authors:** Michael R. Fiorini, Allison A. Dilliott, Sali M.K. Farhan

## Abstract

Traumatic brain injury (TBI), resulting from a violent force that causes functional changes in the brain, is the foremost environmental risk factor for developing dementia. While previous studies have identified specific candidate genes that may instigate worse outcomes following TBI when mutated, TBI-induced changes in gene expression conducive to dementia are critically understudied. Additionally, biological sex seemingly influences TBI outcomes, but the discrepancies in post-TBI gene expression leading to progressive neurodegeneration between the sexes have yet to be investigated. We conducted a whole-genome RNA sequencing analysis of post-mortem brain tissue from the parietal neocortex, temporal neocortex, frontal white matter, and hippocampus of 107 donors characterized by the Aging, Dementia, and Traumatic Brain Injury Project. Our analysis was sex-stratified and compared gene expression patterns between TBI donors and controls, a subset of which presented with dementia. Here we report three candidate gene modules from the female hippocampus whose expression correlated with dementia in female TBI donors. Enrichment analyses revealed that the candidate modules were notably enriched in cardiac processes and the immune-inflammatory response, among other biological processes. In addition, multiple candidate module genes showed a significant positive correlation with hippocampal concentrations of monocyte chemoattractant protein-1 in females with post-TBI dementia, which has been previously described as a potential biomarker for TBI and susceptibility to post-injury dementia. We concurrently examined the expression profiles of these candidate modules in the hippocampus of males with TBI and found no apparent indicator that the identified candidate modules contribute to post-TBI dementia in males. Here, we present the first sex-stratified RNA sequencing analysis of TBI-induced changes to the transcriptome that may be conducive to dementia. This work contributes to our current understanding of the pathophysiological link between TBI and dementia and emphasizes the growing interest in sex as a biological variable affecting TBI outcomes.

## 1 Introduction

A traumatic brain injury (TBI) occurs when the brain jolts violently within the skull due to an external force that damages brain tissue and the surrounding vasculature. (1) TBIs are the primary source of injury associated death, in addition to leading to devastating neuropsychiatric consequences for the survivors of these injuries. (2) Global estimates indicate that as many as 64-74 million people experience a TBI each year. (3) Although the majority of TBI events are considered mild, ∼50% of TBI patients report more than three neuropsychiatric and/or functional symptoms a year post injury. (4) Beyond the personal devastation and the social burden of TBI, the annual financial impact in the US alone is estimated to be $60 billion USD when accounting for direct and indirect costs. (5)

TBIs impair neurological processes that manifest through acute and chronic symptoms, ranging from transient mild cognitive difficulties and neurobehavioral symptoms to devastating physical, cognitive, and psychosocial impairments that pose lifelong consequences. (6) The sum of the neurological damage from TBI depends on an evolving continuum of two sequential stages, namely the primary and secondary insult. (7) The primary insult involves the initial impact that lacerates neuronal cells and the surrounding vasculature. (8) In turn, the secondary insult comprises a vicious interconnected cycle of non-mechanical damage characterized by terminal membrane depolarization, glutamate excitotoxicity, intracellular calcium overload, neuroinflammation, mitochondrial dysfunction, oxidative stress, and neuronal cell necrosis and apoptosis. (8-16) Ultimately, neuronal death leads to tissue loss and, in some cases, neurodegeneration following TBI. (16)

The field of neurotrauma research has been historically biased towards male subjects. (17) Consequently, sex-specific pathologies and long-term impairments of TBI remain unclear. In recent years, however, there has been a greater recognition of the necessity for sex to be investigated as a biological variable affecting clinical outcomes of TBI. While males suffer far more TBIs per year, many patient cohort studies suggest that females experience poorer outcomes following severe brain injury. (2, 18) Although sociological factors contributing to the cause and severity of the TBI may influence these trends, it is hypothesized that genetic factors, comorbidities, extracerebral injuries, and hormonal differences may also impact the observed sex differences. (17)

The magnitude of the prevalence, severity, and complexity of TBI warrants the growing concern for brain injury as the foremost environmental risk factor for neurodegenerative disease. (6) While it now stands that sustaining repeated mild TBIs elevates the risk of developing a pathologically distinct form of dementia known as chronic traumatic encephalopathy, evidence also suggests that a single moderate to severe TBI may be sufficient to trigger Alzheimer’s disease (AD). (19-22) AD is a progressive neurodegenerative disease characterized by chronic dementia and hallmark neuropathological protein depositions, including extracellular amyloid-beta (Aβ) plaques and intracellular neurofibrillary tangles (NFT) composed of hyperphosphorylated tau. (23) Notably, diffuse cortical Aβ plaques accumulate in up to 30% of patients following severe TBI, while in other instances the spatial distribution of NFTs after one episode of severe TBI resembles that of chronic traumatic encephalopathy. (24, 25) Although the contribution of TBI to AD pathophysiology remains poorly understood, it is proposed that the cerebrovascular consequences of TBI may trigger AD pathology, such as Aβ and NFT accumulation.(26) Moreover, the TBI induced Aβ and NFT accumulation may lead to glutamate excitotoxicity, immune inflammation, and proteolytic activity associated with apoptosis exacerbating the aberrant protein depositions and dementia. (9, 27, 28)

Currently, the literature lacks studies investigating post-TBI transcriptional changes that may be conducive to a dementia cascade in TBI patient cohorts. However, animal models have revealed that neuronal cells exhibit altered gene expression following TBI, such that acute gene expression in traumatized tissue moderates the severity of the secondary insult, and therefore the extent of sustained neuronal damage, but persistent expression patterns may trigger and maintain long-term neurodegeneration. (29) In response to the rising concern for TBI as the most robust environmental risk factor for dementia, we conducted a genome-wide RNA sequencing analysis of post-mortem brain tissue from a cohort of donors presenting with and without dementia, of which one subset had experienced TBI (TBI donors), and the other subset had not (controls). Using these data, we identified and characterized sex-specific transcriptome signatures in TBI donors, which may contribute to the pathogenesis of dementia, such as AD.

## 2 Methods

We used R statistical software v4.1.1 in RStudio v4.1.1 to conduct computational analyses and the *ggplot2* R package for data visualization. (30, 31)

### 2.1 Brain sample RNA-Seq data acquisition

We conducted our genome-wide RNA sequencing analysis on the open-source dataset from the Aging, Dementia, and Traumatic Brain Injury Project conducted by the Allen Institute for Brain Science. (32) The dataset included RNA sequencing from post-mortem brains of 107 donors, of which 53 were from individuals that had experienced a TBI (49% presenting with dementia), and 54 were from controls that had not experienced a TBI (44% presenting with dementia). The cohort consisted of 105 self-reported “white” and two “non-white” individuals. An overview of the biological sex, TBI history, and dementia status of the study’s cohort is presented in Table 1 and an overview of the ages at TBI and ages at death of the study’s cohort is presented in Supplementary Table 1. The data included gene expression profiling from four brain regions: 1) the neocortex from the posterior superior temporal gyrus, 2) the neocortex from the inferior parietal lobule, 3) the white matter underlying the parietal neocortex, and 4) the hippocampus. The RNA sequencing methods used on the post-mortem brain tissues were previously described. (33) Briefly, whole transcriptomes were captured using Illumina’s TruSeq Stranded Total RNA Sample Prep Kit, and raw reads were aligned to the GRCh38.p2 human genome. The dataset quantified all RNA sequencing samples by fragments per kilobase of exon per million mapped fragments (FPKM). We further normalized the FPKM data matrix using the zFPKM method described by Hart et *al*. 2013 to retain the biologically relevant genes, setting the active/repressed zFPKM cut-off at -3. (34)

**Table 1.** Sample sizes of the Aging, Dementia, and Traumatic Brain Injury Project cohort based on sample source, TBI status, and dementia status.

| Brain region/Donor category | Dementia | No Dementia | Total |
| --- | --- | --- | --- |
| *Temporal neocortex* |  |  |  |
| Male TBI | 11 | 17 | 28 |
| Male control | 16 | 15 | 31 |
| Female TBI | 14 | 6 | 20 |
| Female control | 7 | 13 | 20 |
| *Parietal neocortex* |  |  |  |
| Male TBI | 10 | 16 | 26 |
| Male control | 15 | 13 | 28 |
| Female TBI | 13 | 4 | 17 |
| Female control | 6 | 14 | 20 |
| *Hippocampus* |  |  |  |
| Male TBI | 9 | 16 | 25 |
| Male control | 13 | 16 | 29 |
| Female TBI | 13 | 6 | 19 |
| Female control | 8 | 14 | 22 |
| *Frontal white matter* |  |  |  |
| Male TBI | 9 | 16 | 25 |
| Male control | 13 | 15 | 28 |
| Female TBI | 13 | 6 | 19 |
| Female control | 8 | 14 | 22 |

Abbreviations: TBI, traumatic brain injury.

### 2.2 TBI-associated differential expression analysis

Using the *limma* R package, we identified TBI-associated differentially expressed genes (DEGs) by conducting differential expression analyses between TBI and control donors separately for each sex and brain region. (35) The *limma* package fits gene-wise linear regression models across all RNA samples to evaluate differential expression by estimating log-ratios between multiple target RNA samples simultaneously. (35) To generate the expression matrix necessary for input into the *limma* pipeline, we log2 transformed the FPKM values, as per author recommendations, when raw counts are unavailable. We predefined the threshold for nominally significant DEGs as p-value < 0.05 and absolute log2 fold change > 0.5.

### 2.3 Weighted gene co-expression network analysis to define gene modules

We conducted a weighted gene co-expression network analysis (WGCNA) of the TBI-associated DEGs, using the *WGCNA* R package, to identify subsets (modules) of highly connected genes. (36, 37) Gene co-expression network analysis is a method to describe the interaction between genes across RNA sequencing samples by assembling highly connected genes into gene modules. The software first creates a similarity co-expression matrix, which measures the concordance between gene expression profiles, by calculating the absolute value of Pearson’s correlation coefficients for all possible gene pairs in the predefined gene set. Then, the co-expression similarity matrix is transformed into an adjacency matrix. Here, we constructed our adjacency matrix using the power adjacency function to create a ‘soft’ adjacency matrix, which ranks all the genes in the network according to their connection strength with a gene under consideration. The power adjacency function requires the specification of an algorithm-computed soft-thresholding power (β) to maintain a scale-free topology fit > 0.9. Finally, the software uses a gene dissimilarity measure coupled with a clustering method to construct gene modules based on high topological overlap, which correspond to the branches of a hierarchical clustering dendrogram and are assigned individual colors. We established the minimum module size, referring to the number of genes included in a module, by setting the dendrogram branch cut-off to a height 0.25 corresponding to a correlation of 0.75. The above sequence of computations was carried out independently for each WGCNA, specifically, stratified by sample brain region and sex.

To identify gene modules that may be involved in development of dementia post-TBI, we calculated the correlation between each gene module’s eigengene and dementia diagnosis amongst our samples. The gene module’s eigengenes were produced by the WGCNAs and act as abstractions representing the expression patterns of all genes assigned to a particular module. (36) The correlation between the eigenvalues and dementia were computed using Pearson’s method and significance was measured at an alpha-level of 0.05. Gene modules approaching significance, defined using an alpha-level of 0.1, were also subject to further investigation if the respective analysis revealed separate gene modules with statistically significant positive correlations to dementia and if the non-significant gene module of interest showed high eigenvalue adjacency (> 0.7) to all statistically significant gene modules. All analyses that did not reveal significant or approaching significant module-trait relationships were excluded from further investigation. In short, gene modules of interest with a correlation to dementia amongst TBI donors are referred to as “candidate modules”.

### 2.4 Candidate module gene set enrichment analyses of gene ontology biological processes

We conducted module-specific gene set enrichment analyses (GSEA) of gene ontology biological processes for all candidate modules using the web-based *Enrichr* tool. (38-40) *Enrichr* takes an unbiased list of genes as input to compute enrichment based on gene-set libraries encompassing existing lists created from prior knowledge. We imported the genes included in each candidate module and submitted our query to retrieve annotations from the *Gene Ontology Biological Process 2021* database. (41, 42) We then visualized the results of our GSEA using the web-based *Enrichr Appyter* tool (v0.2.6). (43)

### 2.5 Defining hub genes and interaction networks

To identify the genes driving the topology of our candidate modules, we identified the top 20 genes with the highest connectivity, hereafter referred to as “hub genes”. First, we imported the node and edge plain text files for each candidate module produced by the WGCNA into the *Cytoscape* software platform (v3.9.1). (44) The node file specified all genes assigned to a gene module, while the edge file described the interaction strengths between genes. To identify the genes with the highest connectivity, we used the *ClosenessCentrality* metric in *Cytoscape*, which referred to the reciprocal of the average shortest path length — defining gene interaction strengths with other genes in the module — and was represented by a number between 0 and 1. For example, the closeness centrality of an isolated gene would have been 0. We mapped the top 20 hub genes to an interaction network using the *Cytoscape* software platform separately for each candidate module.

We aimed to determine whether the expression patterns of the top 20 hub genes from each candidate module were distinct in TBI donors presenting with dementia. To do so, we used the Wilcoxon Rank Sum test to compare the average expression levels of each of the top 20 hub genes between TBI donors presenting with dementia and TBI donors not presenting with dementia stratified by sex to determine if the distinction in expression patterns between TBI donors presenting with and without dementia was sex specific. Significance was measured at an alpha-level of 0.05.

### 2.6 Principal component analyses to define genes driving TBI outcomes

To identify the genes within the candidate modules whose expression patterns distinguish donors according to their TBI history and dementia status, we subjected the FPKM counts of candidate module genes to principal component analysis (PCA) using the *stats* R package. (30) Two independent, sex-stratified, PCAs were conducted to compare the expression patterns of: 1) TBI donors presenting with dementia to controls presenting with dementia, and 2) TBI donors presenting with dementia to TBI donors not presenting with dementia. The percent-contribution of each gene to the primary principal component (PC1) of each PCA was calculated. Then, we identified the genes that were among the top 20 genes with the highest contribution to PC1 in each PCA, subsequently denoted as “genes of interest”, and plotted their eigenvectors using the *factoextra* R package. (45) We measured the expression levels of these genes of interest across all donor categories — 1) TBI presenting with dementia, 2) TBI not presenting with dementia, 3) control presenting with dementia, and 4) control not presenting with dementia — stratified by sex, to determine whether TBI donors presenting with dementia had distinctive expression patterns for these genes of interest and whether differences were sex-specific. We used the Wilcoxon Rank Sum test to compare the expression levels of our genes of interest between all donor categories and measured significance at an alpha-level of 0.05.

### 2.7 Gene expression and protein concentration correlation

The Aging, Dementia, and Traumatic Brain Injury Project dataset quantified molecular changes in protein concentrations for tau, phosphorylated-tau, Aβ species, α-synuclein, inflammatory mediators, and neurotrophic factors using Luminex Assays, reported as a concentration of picogram/nanogram of protein per milligram of tissue, as previously described. (46, 47) We measured the correlation between the FPKM measures of genes of interest — including genes that were major PC1 contributors in both PCAs and primary hub genes— and protein concentrations in the same tissue from which the candidate modules were identified for TBI donors presenting with dementia to infer relevant molecular changes resulting from candidate module expression patterns. The correlation between gene expression and protein concentration was determined using Pearson’s method and significance was measured at an alpha-level of 0.05. Notable gene-protein relationships identified were also analysed in the opposite sex to determine whether the molecular changes were sex-specific. Finally, to determine whether a particular protein that was involved in notable gene-protein relationships could serve as a biomarker for the progression of a dementia cascade following TBI, we conducted sex-stratified comparisons of the protein’s concentration in all four brain regions between TBI donors presenting with and without dementia.

## 3 Results

### 3.1 TBI-associated genes identified using differential expression analysis

Across four brain regions of 107 TBI and control samples, we investigated 17,574 biologically active genes, all of which were subjected to a differential expression analysis between TBI donors and controls, stratified by brain region and sex (Figure 1A). In total, we identified 4,013 nominally significant differentially expressed genes (DEGs) in samples from females and 1,792 nominally significant DEGs in samples from males. Upon stratification by brain region, we again observed more DEGs for each individual brain region in samples from females than from males, and the most DEGs were identified in the white matter underlying the parietal neocortex with 1,473 DEGs in females and 373 in males (Figure 1B).

**Figure 1.**
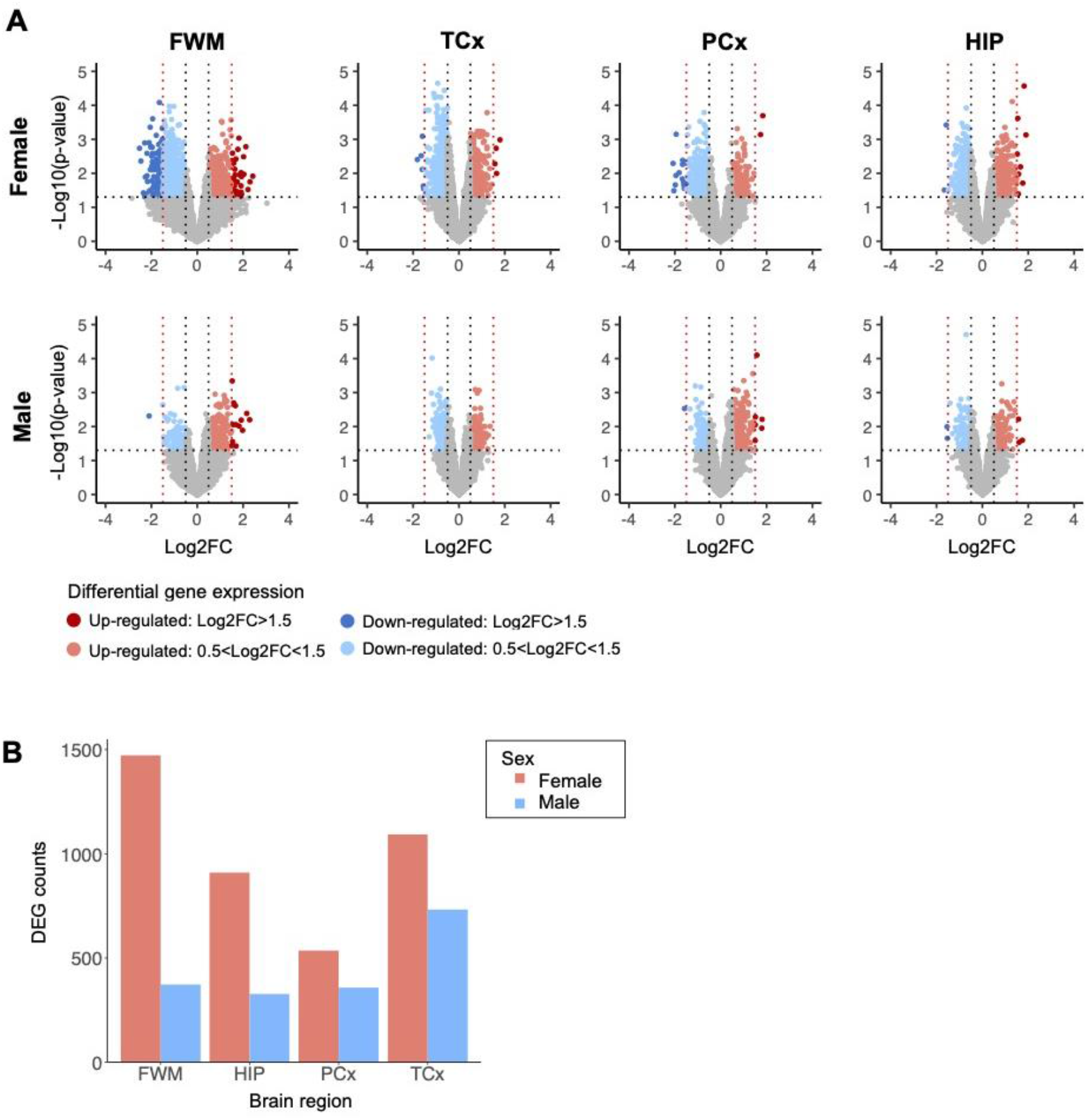
Differential gene expression analysis stratified by brain region and sex. **(A)** Differentially expressed genes (DEGs) identified in TBI donor samples and controls, analyzed independently for each brain region and sex. The threshold for nominal significance was set at p-value < 0.05 and |log2 fold change| > 0.5. The red nodes represent significantly upregulated genes, and the blue nodes represent significantly down-regulated genes. The black and red vertical dotted lines represent a log2 fold change of ±0.5 and ±1.5, respectively. The horizontal dotted line represents a p-value of 0.05. Nominally significant DEGs were subject to independent weighted co-expression network analyses (WGCNA). **(B)** Sum of DEG counts identified in TBI donor samples and controls from each brain region, stratified by sample sex. Abbreviations: DEG, differentially expressed genes; FC, fold change; FWM, frontal white matter; HIP, hippocampus; PCx, parietal neocortex; TCx, temporal neocortex.

### 3.2 Co-expression of TBI-associated DEGs and module-trait relationships

To identify gene modules of highly connected genes, we subjected all nominally significant TBI-associated DEGs (p-value < 0.05 and |log2 fold change| > 0.5) to a weighted gene co-expression network analysis (WGCNA), stratified by brain region and sex. The number of gene modules identified by each independent WGCNA is outlined in Table 2.

**Table 2.** Number of gene modules identified using weighted co-expression network analyses and significant gene modules based on module-trait relationships with dementia status using TBI-associated differentially expressed genes stratified by sex and brain region.

| Brain region | Gene modules |  | Significant gene modules* |  |
| --- | --- | --- | --- | --- |
|  | Male | Female | Male | Female |
| Temporal neocortex | 5 | 12 | 0 | 0 |
| Parietal neocortex | 3 | 3 | 0 | 0 |
| Hippocampus | 3 | 8 | 0 | 2 |
| Frontal white matter | 4 | 8 | 0 | 0 |

\* Statistically significant correlation ( $p < 0.05$ ) between TBI donor expression patterns and dementia status.
Analyses were conducted using Pearson's method at an alpha level of 0.05.
Abbreviations: TBI, traumatic brain injury.

We used Pearson’s method to identify correlations between the gene modules and dementia following TBI. The only gene modules that were significantly associated with the development of dementia following TBI were from female hippocampal samples. Specifically, 910 TBI-associated DEGs identified from the hippocampus of 19 female TBI donors were used to construct the co-expression network. A soft thresholding power (β) of four was used to maintain an approximate scale-free topology with a scale-free topology fit > 0.9 (Figure 2A). WGCNA of the female hippocampus identified eight distinct gene modules which varied in size from 13 genes (grey module) to 390 genes (turquoise module) (Figures 2B-C). A network heatmap defining the topological overlap of all genes assigned to the gene modules was constructed and indicated a moderate level of overlap between gene modules (Figure 2D). Comparison of the eigenvalues of each gene module using a hierarchical clustering dendrogram and an adjacency heat map showed high relatedness between the red, black, and green gene modules and between the brown and turquoise gene modules (Figure 2E). Upon examination of the module-trait relationship between each gene module from the female hippocampus and dementia status amongst female TBI donors (n = 19) (Figure 2F), the black module (Supplementary Table 2) and green module (Supplementary Table 3) displayed significant positive correlations to dementia (r = 0.57, p = 0.01 and r = 0.55, p = 0.01, respectively). Additionally, the red module (Supplementary Table 4) displayed a positive correlation that approached significance (r = 0.4, p = 0.09) with strong adjacency (> 0.7) to both the black and green modules and was therefore included in downstream analyses.

**Figure 2.**
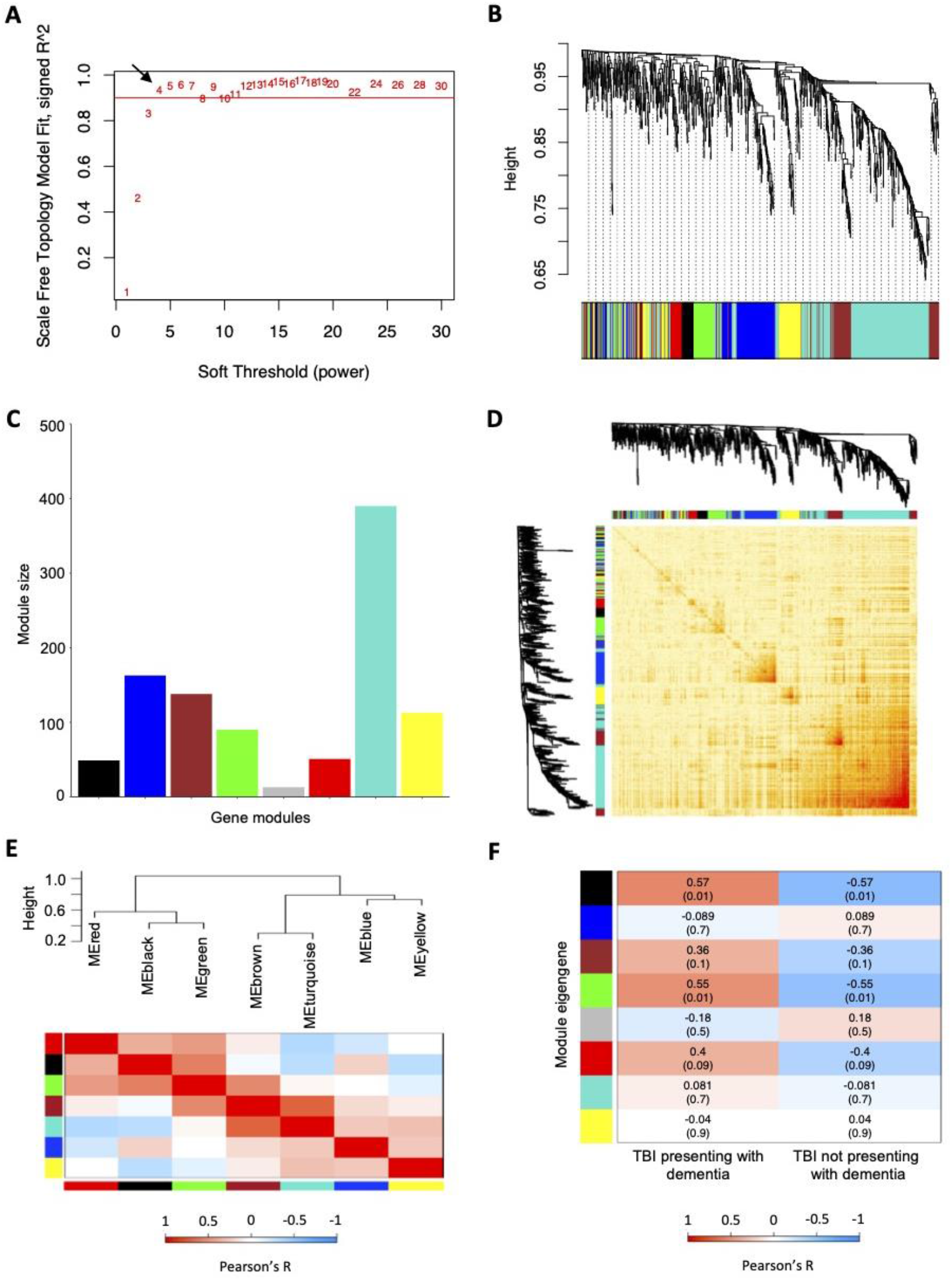
Weighted co-expression network analysis for TBI-associated differentially expressed genes identified from the female hippocampus. **(A)** Scale free fitting index of different soft thresholds (β), showing the scale-free model fit as a function of β. Four was selected as the β for the female hippocampus analysis. **(B)** Co-expression cluster dendrogram of TBI-associated differentially expressed genes (DEGs). Gene modules were assigned unique colors. Each line represents a single gene, and the height on the y-axis represents dissimilarity in expression. **(C)** Number of genes included in each computed co-expression modules. Node colors correspond to assigned module colors. **(D)** Network heat map displaying the topological overlap of all TBI-associated DEGs assigned to each co-expression module. The co-expression dendrogram and assigned module colors are shown along the X and Y axes. Light yellow indicates low topological overlap, and dark red indicates high topological overlap. **(E)** Eigengene hierarchical clustering dendrogram and adjacency heatmap of modules eigengenes (ME). In the dendrogram, branches represent module eigengenes, and positively correlated eigengenes are grouped. In the heatmap, each row and column correspond to a specific module’s eigengene, denoted by module color. **(F)** Module-trait relationships analyzed using Pearson’s method to assess the correlation between dementia status in TBI patients and each module’s eigengene. Correlation coefficients (p-values) are indicated in each respective cell. Abbreviations: ME, module eigengene; TBI, traumatic brain injury.

No additional gene modules were significantly positively correlated with dementia in female TBI donor samples from the remaining brain regions, nor in male TBI donor samples from any brain regions (Supplementary Figure 1). Accordingly, subsequent analyses only encompassed candidate modules from the female hippocampus.

### 3.3 Enrichment of biological processes in candidate module genes

Module-specific GSEAs were conducted for each candidate module from the female hippocampus to identify functional neurodegenerative mechanisms that may precipitate dementia post-TBI. The genes from the black module were annotated to 190 gene ontology biological processes, of which four were significantly enriched (p-value <0.05) following Benjamin-Hochberg multiple testing correction, including RNA pseudouridine synthesis and modification (Figure 3B). The genes from the green module were annotated to 441 gene ontology biological processes, of which zero were significantly enriched following multiple testing correction; however, 60 biological processes were significantly enriched prior to multiple testing correction, including glycosylation, inflammatory response, alpha-beta T cell differentiation, and cardiac muscle contraction and membrane repolarization (Figure 3D). Notably, cardiac-associated processes were significantly enriched in both the black and green modules prior to correction for multiple testing. Lastly, the red module was annotated to 457 gene ontology biological processes, of which 14 were significantly enriched following multiple testing correction, including the immune-inflammatory response, regulation of major histocompatibility complex class II biosynthetic processes and regulation of monocyte chemotactic protein-1 (MCP-1) production (Figure 3F).

**Figure 3.**
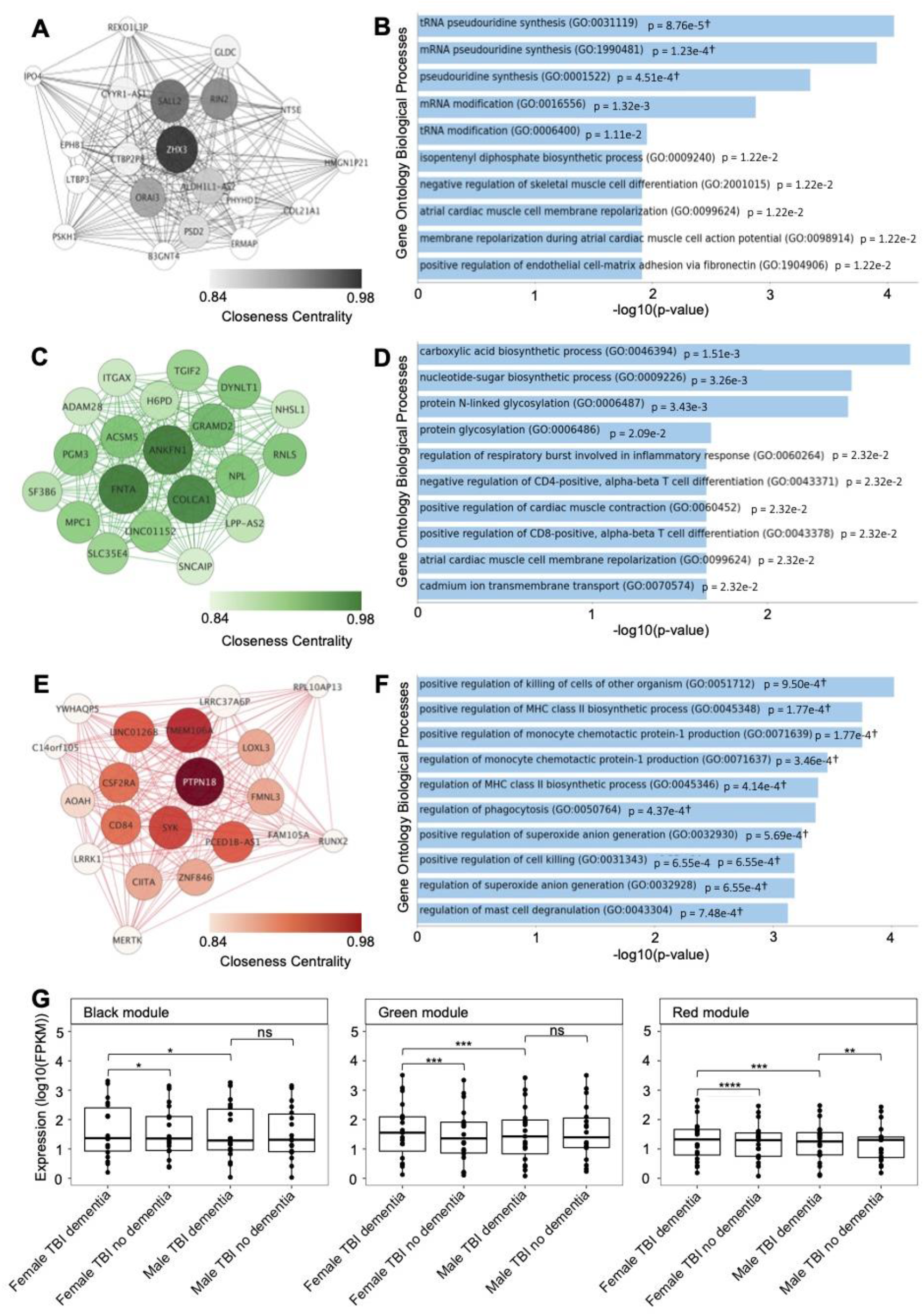
Top hub genes and module-specific gene set enrichment analysis (GSEA) of candidate modules from the female hippocampus. **(A)** Interaction networks of the top 20 genes with the highest connectivity (hub genes) in the black candidate module, identified in the weighted gene co-expression network analysis (WGCNA) of the female hippocampus. (**B)** The top 10 enriched Gene Ontology Biological Processes for the black candidate module, identified using GSEA. **(C)** Interaction networks of the top 20 genes with the highest connectivity (hub genes) in the green candidate module, identified in the weighted gene co-expression network analysis (WGCNA) of the female hippocampus. (**D)** The top 10 enriched Gene Ontology Biological Processes for the green candidate module, identified using GSEA. **(E)** Interaction networks of the top 20 genes with the highest connectivity (hub genes) in the red candidate module, identified in the weighted gene co-expression network analysis (WGCNA) of the female hippocampus. Interaction networks were created using *Cytoscape* (version 3.9.1). (**F)** The top 10 enriched Gene Ontology Biological Processes for the red candidate module, identified using GSEA. †Overlap of less than five genes. **(G)** Comparison of the expression — fragments per kilobase of transcript per million mapped reads (FPKM) — of the top 20 hub genes from each candidate modules between samples from female TBI donors with dementia compared with female TBI donors without dementia, and male TBI donors with dementia using the Wilcoxon’s Rank Sum test. Expression levels for male TBI donors with dementia were also compared to male TBI donors without dementia. * p < 0.05, ** p < 0.01, *** p < 0.001, **** p < 0.0001, ns = not significant. Abbreviations: FPKM, fragments per kilobase of transcript per million mapped reads; TBI, traumatic brain injury.

### 3.4 Identification of candidate module hub genes

We identified the top 20 hub genes for the black, green, and red modules from the female hippocampus using the *Cytoscape ClosenessCentrality* metric. The hub gene network of the black module (n = 49 genes) contained 182 gene-gene interaction edges, of which *ORAI3, RIN2, SALL2*, and *ZHX3* had closeness centrality measures > 0.9 (Figure 3A). The green module (n = 90 genes) contained 182 gene-gene interaction edges, of which *ACSM5, ANKFN1, COLCA1, DYNLT1, FNTA, GRAMD2, LINC01152, MPC1, NPL, PGM3*, and *RNLS* had closeness centrality measures > 0.9 (Figure 3C). Finally, the red module (n = 51 genes) contained 185 gene-gene interaction edges, of which *CD84, CSF2RA, LINC01268, PCED1B-AS1, PTPN18, SYK*, and *TMEM106A* had closeness centrality measures > 0.9 (Figure 3E).

Interestingly, when we compared the hippocampus expression patterns of the candidate modules top 20 hub genes female TBI donors presenting with dementia displayed significantly higher expression patterns for all three candidate modules compared to female TBI donors not presenting with dementia, including the black module (p = 1.3e-2), green module (p = 3.3e-4), and red module (p = 7.2e-5) (Figure 3G). However, upon repetition of the analysis using male TBI hippocampal samples only the top 20 hub genes from the red module displayed a significant difference in gene expression between donors presenting with and without dementia (p = 1.2e-2), with TBI donors presenting with dementia showing higher expression levels. Finally, we compared expression patterns of the top 20 hub genes from each candidate module between male and female TBI donors presenting with dementia and found significantly higher expression levels in female samples compared to male samples from all modules, including the black module (p = 3.8e-2), green module (p = 6.4e-4), and red module (p = 2.6e-4).

### 3.5 Principal component analyses to identify genes driving TBI outcomes

We conducted two independent PCAs to identify genes within the candidate modules from the female hippocampus that successfully characterize female samples according to their TBI history and dementia status. The first PCA incorporated fragments per kilobase of exon per million mapped fragments (FPKM) for all candidate module genes from hippocampal samples of female TBI donors presenting with dementia and female controls presenting with dementia, highlighting TBI-associated distinctions in module expression patterns (Figure 4A). The primary and secondary PCs explained 25% and 14.75% of the variation, respectively. The second PCA incorporated all candidate module gene FPKM counts from hippocampal samples of female TBI donors presenting with dementia and female TBI donors not presenting with dementia to identify dementia-associated distinctions in expression patterns (Figure 4B). The primary and secondary PCs explained 27.04% and 10.9% of the variation, respectively. In both PCAs we found a clear distinction along PC1 between the respective categories of female donors, suggesting that the transcriptional landscape of the candidate modules may distinguish female donors according to both their history of TBI and dementia status.

**Figure 4.**
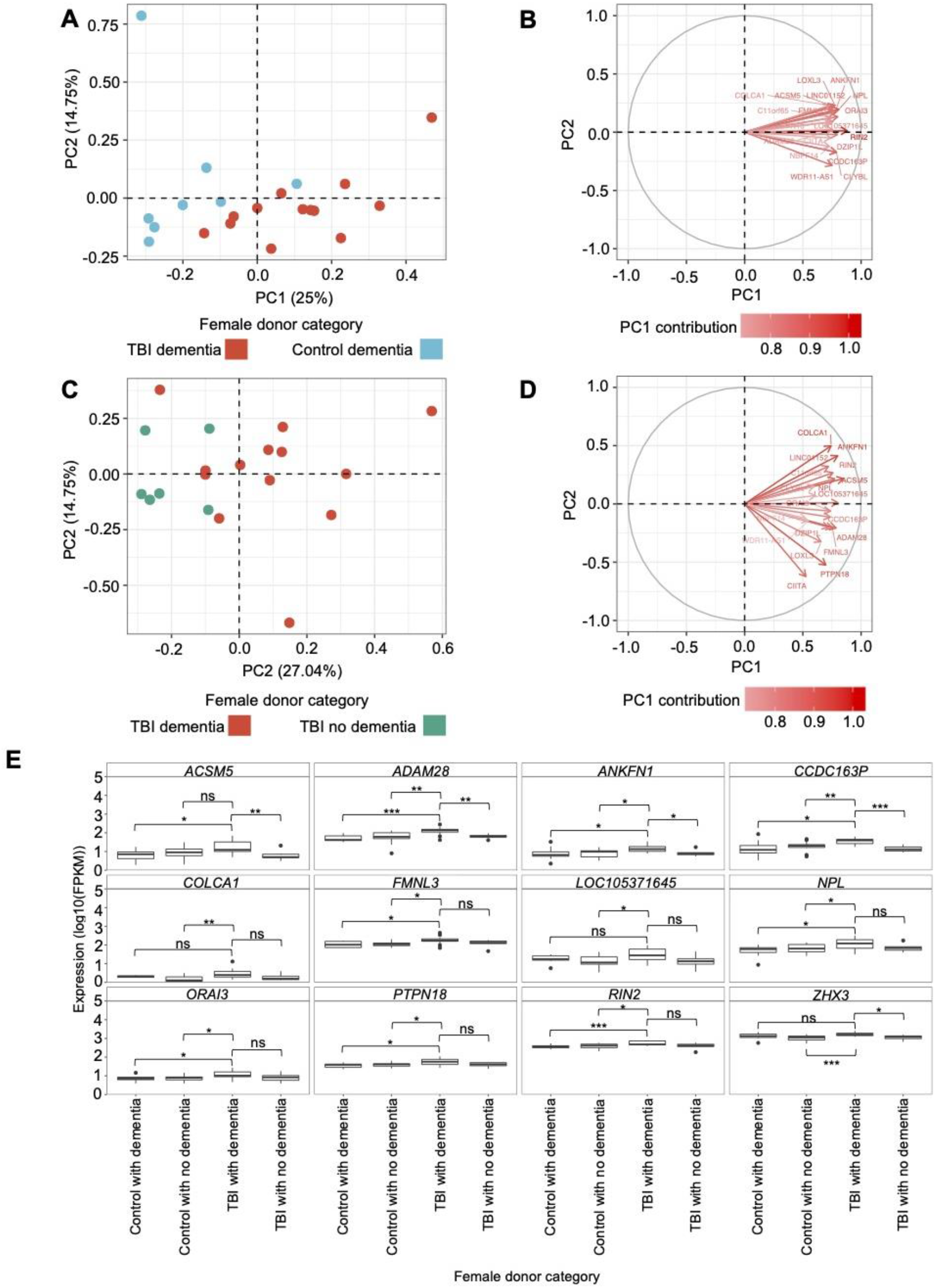
Principal component analysis (PCA) and gene-protein correlations of candidate modules from the female hippocampus. **(A)** PCA using fragments per kilobase of transcript per million mapped reads (FPKM) of the genes assigned to candidate modules in the hippocampus of female TBI donors presenting with dementia and female controls presenting with dementia. The primary and secondary PCs are shown, and their contribution to the variation is indicated on the respective axis. **(B)** Eigenvectors and percent contribution of the top 20 genes showing the highest contribution to PC1 in plot A, which compared the expression of candidate module genes in the hippocampus between TBI donors presenting with dementia and controls presenting with dementia. Progressively darker red color indicates higher contribution to PC1. **(C)** PCA using FPKM of the genes assigned to candidate modules in the hippocampus of female TBI donors presenting with dementia and female TBI donors not presenting with dementia. **(D)** Eigenvectors and percent contribution of the top 20 genes showing the highest contribution to PC1 in plot C, which compared the expression of candidate module genes in the hippocampus between TBI donors presenting with dementia and TBI donors not presenting with dementia. **(E)** Expression levels of genes of interest — including primary hub genes (*ANKFN1, PRPN18, ZHX3*) and genes that were amongst the top 20 contributors to PC1 in both PCAs — in the female hippocampus were compared using the Wilcoxon Rank Sum test between female TBI donors with dementia and all other categories of female donors. Within each panel, boxplots for each category of female donor are displayed in the same order as the figure legend. * p < 0.05, ** p < 0.01, *** p < 0.001, ns = not significant. Abbreviations: FPKM, fragments per kilobase of transcript per million mapped reads; PC1; primary principal component; PC2, secondary principal component; TBI, traumatic brain injury.

We computed the percent contribution of all candidate module genes to PC1 from both PCAs. Among the top 20 contributing genes to each PC1, we found 10 genes overlapping between the two PCAs — *ACSM5, ADAM28, ANKFN1, CCDC163P, COLCA1, FMNL3, LOC105371645, NPL, ORAI3*, and *RIN2* (Figures 4C-D) — which, along with the primary hub gene from each candidate module (*ANKFN1, PTPN18, ZHX3*), are hereafter referred to as “genes of interest”. Using the Wilcoxon Rank Sum test, we compared the hippocampal expression levels of the genes of interest in the female TBI donors presenting with dementia to: 1) female TBI donors not presenting with dementia, 2) female controls presenting with dementia, and 3) female controls not presenting with dementia (Figure 4E; Supplementary Table 5). Female TBI donors presenting with dementia, on average, showed higher expression levels for all genes of interest compared to all other categories of female donors in the dataset, with 27 of the 36 comparisons meeting statistical significance.

Using the genes assigned to candidate modules from the female hippocampus, we again performed two PCAs using the hippocampal samples from male donors to determine whether the contributions of the top 20 genes from the candidate modules to TBI and dementia status were sex specific. Here, the PCAs did not indicate a distinction between male TBI donors presenting with dementia and male controls presenting with dementia, nor between male TBI donors presenting with dementia and male TBI donors not presenting with dementia (Supplementary Figure 2). Finally, we again used the Wilcoxon Rank Sum test to compare the hippocampal expression levels of the genes of interest in the male TBI donors presenting with dementia to: 1) male TBI donors not presenting with dementia, 2) male controls presenting with dementia, and 3) male controls not presenting with dementia (Supplementary Figure 2E; Supplementary Table 5). The only gene demonstrating differential expression was *CCDC163P*, which was expressed significantly higher in male TBI donors presenting with dementia compared to male controls not presenting with dementia (p = 4.3e-2).

### 3.6 Correlation of gene of interest expression to notable protein concentrations

We computed the correlation between the FPKM measures of our genes of interest and protein concentrations to determine whether expression level may be influencing dementia-related pathological markers in the hippocampus of female TBI donors (Figure 5A). We found that MCP-1 concentration was significantly positively correlated with expression levels of *ACSM5* (r = 0.7, p = 1.1e-2), *ADAM28* (r = 0.64, p = 2.4e-2), *ANKFN1* (r = 0.89, p = 0.1e-3), *COLCA1* (r = 0.9, p = 5.4e-5), *LOC105371645* (r = 0.81, p = 1.5e-3), *NPL* (r = 0.77, p = 3.2e-3), and *RIN2* (r = 0.59, p = 4.2e-2). Additionally, *NPL* was significantly negatively correlated with IL7 concentration (r = -0.62, p = 3.2e-2), *LOC105371645* was significantly negatively correlated with IL6 concentration (r = -0.66, p = 1.9e-2), and *PTPN18* was significantly negatively correlated with phosphorylated tau concentration (r = -0.79, p = 2.4e-3), yet significantly positively correlated with tau concentration (r = 0.65, p = 2.2e-2).

**Figure 5.**
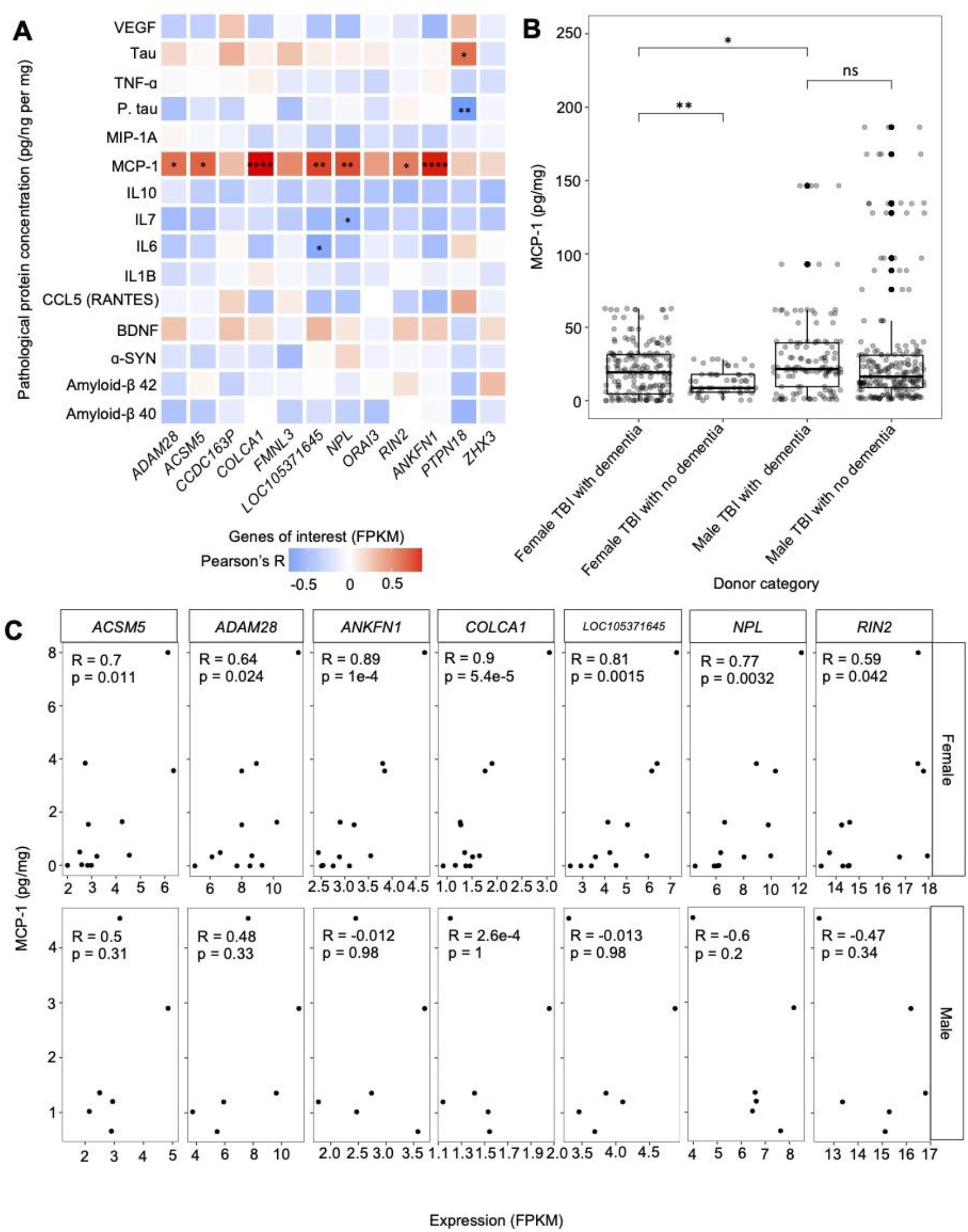
Correlation between the expression of the genes of interest and protein concentrations. **(A)** Gene-protein correlation computed using Pearson’s method between the genes of interest — including primary hub genes (*ANKFN1, PRPN18, ZHX3*) and genes that were amongst the top 20 contributors to PC1 in both PCAs; comparing the expression levels of all candidate module genes in the hippocampus of female TBI donors presenting with dementia to female controls presenting with dementia and female TBI donors not presenting with dementia— fragments per kilobase of transcript per million mapped reads (FPKM) and protein concentration (pg/ng per mg) in the hippocampus of female TBI donors presenting with dementia. **(B)** Comparison of the concentration of hippocampal MCP-1 between female TBI donors presenting with dementia and female TBI donors not presenting with dementia, male TBI donors presenting with dementia using the Wilcoxon’s Rank Sum test. MCP-1 concentrations for male TBI donors presenting with dementia were also compared to male TBI donors not presenting with dementia. **(C)** Correlation computed using Pearson’s method between genes of interest expression (FPKM) and MCP-1 concentration (pg/mg) for all statistically significant relationships. Pearson correlation coefficients and p-values are displayed in each plot. * p < 0.05, ** p < 0.01, *** p < 0.001, **** p < 0.0001, ns = not significant. Abbreviations: FPKM, fragments per kilobase of transcript per million mapped reads; TBI, traumatic brain injury.

Given that 7 of the 12 genes of interest from the candidate modules identified in the female hippocampus showed a statistically significant positive correlation with MCP-1 concentrations in hippocampal tissue of females with TBI presenting with dementia, we further analysed these gene-protein relationships in the hippocampus of male TBI donors presenting with dementia (Figure 5B). Interestingly, we not only failed to replicate the significant gene-protein relationships within our male samples, but 3 of the 7 genes showed negative correlations with MCP-1 concentrations, namely *LOC105371645, NPL*, and *RIN2*, albeit these relationships were non-significant. Finally, we investigated the aggregate concentration of MCP-1 in all brain regions of female and male TBI donors presenting both with and without dementia to evaluate the potential of MCP-1 to serve as a biomarker for the progression of a dementia cascade following TBI. We found that samples of female TBI donors presenting with dementia had significantly higher concentrations of MCP-1 compared to female TBI donors not presenting with dementia (p = 5.6e-3) (Figure 5C). In contrast, no significant difference in MCP-1 concentration was observed between male TBI donors presenting with and without dementia. Notably, male TBI donors presenting with dementia showed higher concentrations of MCP-1 compared to female TBI donors presenting with dementia (p = 0.02).

## 4 Discussion

We conducted a sex-stratified whole-genome RNA sequencing analysis of post-mortem brain tissue from the parietal neocortex, temporal neocortex, frontal white matter, and hippocampus and compared gene expression patterns between TBI donors and controls, some of whom presented with dementia, to further our understanding of how TBI may precipitate the development of a dementia cascade. Although we did not identify any significant relationships between gene expression and post-TBI dementia in our male samples, we report three candidate modules of co-expressed genes from the female hippocampus that correlated with dementia among our female TBI samples. Gene set enrichment analysis (GSEA) revealed that these candidate modules were notably enriched in cardiac processes and the immune-inflammatory response among other biological processes. Upon subjecting the candidate module genes from the female hippocampus to PCA, we identified ten genes of interest that distinguish the expression patterns of female TBI donors presenting with dementia according to their TBI history and dementia status and may serve as therapeutic targets to moderate the progression of a dementia cascade following TBI. In addition, most of these genes of interest, including the primary hub genes from each candidate module, showed a significant positive correlation with monocyte chemotactic protein-1 (MCP-1) concentration in the female hippocampus, a potential biomarker for susceptibility and progression of post-TBI dementia. We concurrently examined the expression profiles of these candidate modules in the hippocampus of male samples and found no apparent indicator that the identified genes contribute to post-TBI dementia in males, suggesting a sex dependence in the TBI-induced transcriptional mechanisms involved in dementia cascades following TBI.

Our results suggest that hippocampal expression patterns play prominent roles in the development of post-TBI dementia in females. The hippocampus is often a critical determinant of TBI outcomes due to its prominent role in learning and memory and its high susceptibility to neurotrauma. (48) In fact, prolonged cognitive deficits in animal models have resulted from TBI-induced dysfunction in hippocampal circuits, including changes in cell excitability, slower axon conduction velocity, and impaired long-term potentiation. (49-54) Past studies have identified acute transcriptional changes following TBI in hippocampal tissue associated with oxidative stress, metabolism, inflammation, structural changes, and cellular signaling. (55) While we also identified several of these biological process in the GSEA of our hippocampal candidate modules, they did not meet significance after adjustment for multiple testing. Instead, our analyses revealed significant enrichments in the immune inflammatory response. In addition, cardiac processes were enriched in two of the three candidate modules, and although these results were only found to be significant prior to multiple testing correction, the convergence of the biological processes from to different candidate modules warrants further discussion.

In accordance with our observation that impaired cardiac processes may drive a dementia cascade in females post-TBI, systolic myocardial dysfunction is common in moderate-severe TBI sequalae due to a combination of catecholamine excess and systemic inflammatory response. (56) Among other pathological consequences, myocardial dysfunction contributes to cerebral hypoperfusion which drives a neuronal energy crisis and may instigate defects in protein synthesis that lead to the deposition of amyloid-beta (Aβ) plaques and neurofibrillary tangles (NFT), hallmarks of Alzheimer’s disease (AD). (56, 57) Given the well documented heart-brain continuum, coupled with our results which show that genes involved in atrial contraction and atrial membrane repolarization have altered expression in females presenting with dementia post-TBI, it seems plausible that the prolonged TBI-induced impairment of cardiac processes drive a dementia cascade following head injury. Furthermore, cerebral perfusion decreases naturally with increasing age, which, when exacerbated by TBI, may explain the higher incidence of post-TBI dementia amongst females in our dataset, whose mean age at injury exceeded male counterparts by 25 years.

GSEA of the candidate modules from the female hippocampus also showed enrichments in immune/inflammation processes, including interleukin 6, tumor necrosis factor-alpha, and major histocompatibility complex class II production. TBI induced neuroinflammation comprises both neuroprotective and neurotoxic components, the latter includes immune-mediated atrophy, neuronal loss, and axonal degeneration. (58) Furthermore, neuroinflammatory proteins have shown an association with Aβ plaque deposition in AD patients. (59) Interleukin 6 and tumor necrosis factor-alpha are pro-inflammatory, microglia-activating cytokines which have been previously shown to upregulate immediately upon tissue damage from TBI. (60, 61) Similarly, severe TBI patients have shown abnormally high levels of major histocompatibility complex class II molecules on activated microglia. (62, 63) Taken together, these results suggest elevated levels of activated microglia in the hippocampus of female TBI donors presenting with dementia and support previous work which proposed a pathological link between TBI and dementia in that resident microglia are the key cells mediating the inflammatory processes in both AD and in long-term survivors of TBI. As our analyses were conducted on post-mortem brains, on average 29.3 years after our female samples sustained TBI, we cannot comment on the acute inflammatory response of head injury; however, the long-term elevated inflammatory response which we observed in our analyses may be explained by the previously described cytokine cycle, whereby TBI initiates a neuroprotective inflammatory response which may evolve to become self-sustaining and contribute to long-term neurodegenerative changes. (64) In addition, given that post-TBI cytokines — such as interleukin 6 and tumor necrosis factor-alpha — may trigger a self-perpetuating cytokine cycle, and that there is a documented association between inflammation and cardiac dysfunction as discussed previous, we hypothesize that prolonged-inflammation and the consequent cardiac abnormalities may conspire to bring about dementia pathologies in certain females with TBI.

In addition to the inflammatory processes identified in the GSEA, the production of monocyte chemotactic protein-1 (MCP-1) was significantly enriched in the candidate modules. MCP-1 is involved in macrophage recruitment into damaged tissue, but elevated expression has been shown to degrade the axonal myelin sheath, (65-69) and previous research has suggested that the post-TBI induction of MCP-1 may impose higher risk of AD due to the demyelination process that reduces the brain’s resilience to subsequent neurodegenerative insults. (70) Interestingly, seven of the ten genes of interest identified in our PCA — including *NPL, LOC105371645, ADAM28, COLCA1, ACSM5, RIN2*, and *ANKFN1* — showed statistically significant positive correlations with MCP-1 concentrations in the hippocampus of female TBI donors presenting with dementia. Moreover, female TBI donors presenting with dementia had statistically significant higher concentrations of MCP-1 in the hippocampus compared to female TBI donors not presenting with dementia. The distinct expression patterns of our genes of interest and their associations with MCP-1 concentrations in the hippocampus of female TBI donors presenting with dementia allow us to support the previous proposition that MCP-1 may, with future research, serve as a molecular biomarker for TBI and susceptibility to post-injury AD, particularly in females. (70)

Additionally, the genes of interest distinguished the expressional landscape of hippocampal tissue from females with TBI that presented with dementia may serve as therapeutic targets to moderate transcriptional changes contributing to the progression of a TBI-induced dementia cascade. More specifically, we found that ten genes of interest — *ACSM5, ADAM28, ANKFN1, CCDC163P, COLCA1, FMNL3, LOC105371645, NPL, ORAI3*, and *RIN2* — that distinguished the transcriptional patterns in the hippocampus of female TBI donors presenting with dementia compared to controls presenting with dementia and to TBI samples not presenting with dementia. Among these genes, *ACSM5, ANKFN1, COLCA1, NPL, ORAI3*, and *RIN2* demonstrated a closeness centrality > 0.9 in their respective gene modules and therefore were largely responsible for defining the transcriptional topology, or expression patterns, of the modules being investigated to drive post-TBI dementia in females. Further, female TBI samples with dementia had higher expression of all genes of interest compared to all other categories of female donors. Thus, these genes describe quantifiable distinctions in the expressional landscape of the hippocampus of female TBI donors presenting with dementia and offer a prospective avenue to regulate the progression of, and susceptibility to, post-TBI dementia in female patients. While our samples consist of post-mortem tissues that fail to depict the temporal fluctuations in the gene’s expressional patterns, this shortcoming could be mediated through animal model RNA sequencing analyses that monitor the expression levels of these genes over prolonged periods to resolve their time-dependent expressional patterns and their association with neurodegeneration.

A primary goal of our analyses was to describe the discrepancy between the sexes in the TBI-induced transcriptional changes that lead to a dementia cascade, as past studies using animal models have revealed sex-dependent differences in the pathological and neurological outcome following TBI. (71) Unfortunately, we did not identify any candidate modules from our male samples and therefore could not make direct, inter-sex comparisons of TBI-induced neurodegenerative mechanisms. Instead, we evaluated the transcriptional patterns of the candidate modules identified from the female hippocampus in the male hippocampus, and we observed that male TBI donors presenting with dementia had higher expression levels of the top 20 hub genes from the red modules compared to male TBI donors not presenting with dementia. The observation was unsurprising, as the red module was enriched in immune-inflammatory processes, which have been repeatedly cited as major components of post-TBI neurodegeneration. In contrast, there was no difference in the expression of the top hub genes from the black module among male TBI donors according to dementia status, potentially suggesting that changes in cardiac processes may not drive post-TBI dementia in males, or instead may not have as large of a role.

Similarly, the expression patterns of the candidate modules in the hippocampus of male TBI donors presenting with dementia were indistinguishable from hippocampal samples from other male donor categories when subjected to PCA and when expression levels of our genes of interest were compared. Finally, we found no significant associations between the hippocampal expression of our genes of interest and MCP-1 concentrations in the hippocampus of male TBI donors presenting with dementia. Taken together, these results support our hypothesis that there are sex-specific transcriptional mechanisms driving post-TBI sequalae and susceptibility to dementia; however, the driving mechanisms behind our findings require further exploration and validation by independent techniques.

One possibility is that sex hormones may drive the discrepancy in TBI-induced gene expression that we observed between the sexes. Specifically, female sex hormones may provide endogenous neuroprotection in females during their reproductive years as postpubescent teenage females generally experience better outcomes following TBI than age-matched males. (72) Accordingly, animal studies have found that female sex hormones improve outcomes post-TBI, yet clinical trials involving the administration of progesterone post-TBI did not show improvement of outcome. (17, 73, 74) But it is important to recognize the normal depletion of sex hormones in the ageing process. For women, menopause initiates a relatively rapid loss of estradiol and progesterone, while ageing men experience a relatively gradual loss of testosterone. (75) Postmenopausal females have demonstrated higher mortality rates following TBI than age-matched males, which again may suggest a neuroprotective role of female gonadal hormones. (76) Regarding dementia, the accompanying hormonal changes of menopause may accelerate the development of metabolic and cardiovascular disease which are well documented risk-factors for late-life dementia. (77-79) In our dataset, the average age at TBI was 56 years for females, while menopause typically occurs at 55 years of age. Therefore, the average postmenopausal age at injury for our female samples, and the consequential hormonal depletion, may have contributed to the transcriptional changes that we found were associated with dementia in female donors. Furthermore, due to our findings that cardiovascular processes may be involved in the dementia cascade in female TBI donors, we propose the possibility that TBI may exacerbate the naturally occurring heightened risk for cardiovascular disease in postmenopausal female; therefore, increasing susceptibility to dementia.

Average age at injury for males may in part, be responsible for the lack of candidate modules showing a significant association to dementia. The average age at injury for males in this dataset was 31 years, 25 years younger than our female samples and substantially lower than 55 years old, which has been previously described as a benchmark age beyond which the risk for dementia after TBI increases significantly. (80) The overall interaction between age, TBI, and dementia suggests that individuals who suffer brain injury at a younger age may be more resilient to the long-term neurodegenerative effects. (80) Accordingly, the possibility exists that the lack of associations between transcriptional changes and post-TBI dementia in male samples was due to a relatively young average age at injury. Contrarily, the average age of injury for our female donors surpassed the cut-off for elevated risk to dementia. To further our understanding of the disparity in post-TBI susceptibility to dementia between the sexes, analyses should be repeated using age matched cohorts for females and males.

Further, we must recognize that TBI-induced changes in gene expression may be more subtle in men and therefore may require larger sample sizes to detect. Our initial differential expression analysis identified fewer nominally significant differentially expressed genes (DEG) for each brain region among males according to their TBI history than females, introducing the possibility that less extensive changes in TBI-induced gene expression led to the exclusion of relevant genes from our weighted gene co-expression network analysis (WGCNA) of male samples. Nonetheless, our male TBI donor sample sizes were greater than that of female TBI donors, suggesting that females may experience greater consequences to TBI at a transcriptomic level resulting in greater susceptibility to poor outcomes following brain injury.

We recognize that our study has notable limitations. Firstly, our sample size for each analysis was relatively small, given our need to subset the dataset according to biological sex, brain region, TBI history, and dementia status. Yet, this work is the first RNA sequencing analysis to investigate sex-specific TBI-induced transcriptional changes that may lead to the development of dementia, and our results highlight the importance for further analyses with larger datasets. Further, ancestral background was not genetically ascertained within our analyses, although self-reported ethnicities were documented. We also did not identify candidate modules from our male samples, resulting in the inability to compare TBI-induced neurodegenerative mechanisms between the sexes. Nonetheless, we aimed to see if the candidate modules from the female hippocampus showed an association with dementia in male TBI hippocampal samples and did not observe any replication between the sexes. Finally, our results need to be validated using a secondary RNAseq cohort, to ensure our observations were not spurious, and require future validation via transcriptomic validation analyses to resolve how the candidate modules contribute to neurodegeneration following TBI.

The exact mechanisms involved in the development of dementia pathologies following TBI remain unclear. Even more uncertain is how TBI-induced neurodegenerative processes differ between the sexes. We found that expression patterns in the female hippocampus associated with cardiac and immune-inflammatory processes showed an association with post-TBI dementia in females, and that these expression patterns did not show an association with dementia in males. In addition, we identified ten female-specific genes of interest that could potentially serve as therapeutic targets, and MCP-1 concentrations in the female hippocampus which could serve as a biomarker to define the progression of a dementia cascade following TBI. To our knowledge, this study is the first sex-stratified RNA sequencing analysis on a human cohort that investigated how TBI-induced changes to the transcriptome may lead to the development of dementia.

## Supporting information

Author Disclosures

STROBE Checklist

## Data Availability

All analysed data are available online at https://aging.brain-map.org/overview/home

## 5 Data availability statement

The datasets for this study can be found online from the Aging, Dementia, and TBI Study conducted by the Allen Institute for Brain Science. https://aging.brain-map.org/overview/home

## 6 Funding

Sali M.K. Farhan is supported by grants from Brain Canada, ALS Canada, and the Tanenbaum Open Science Institute (TOSI) at The Neuro, McGill University.

## 7 Abbreviations

Aβ: amyloid-beta
AD: Alzheimer’s disease
β: soft-thresholding power
DEG: differentially expressed genes
FPKM: fragments per kilobase of exon per million mapped fragments
GSEA: gene set enrichment analysis
MCP-1: monocyte chemotactic protein-1
NFT: neurofibrillary tangles
PCA: principal component analysis
PC1: primary principal component
TBI: traumatic brain injury
WGCNA: weighted gene co-expression network analysis

## 8 Acknowledgments

We want to acknowledge the consent and participation of all individuals from which post-mortem brain analyses were conducted. Further, we thank the Allen Institute for Brain Science and their work in assembling the Aging, Dementia, and TBI Study datasets. A.A.D. is supported by the Canadian Institute of Health Research Banting Postdoctoral Fellowship program. We also thank Dr. Jacqueline Bede and the Departments of Plant Science and Biology at McGill University.

## Notes

### Competing Interest Statement

The authors have declared no competing interest.

### Funding Statement

This study did not receive any funding.

### Author Declarations

The datasets for this study are openly available and can be found online from the Aging, Dementia, and TBI Study conducted by the Allen Institute for Brain Science. https://aging.brain-map.org/overview/home

