## Supplementary Tables 1, 2, 3, 4, 5 and Supplementary Figures 1 and 2 for "Sex-stratified RNA-seq analysis reveals traumatic brain injury-induced transcriptional changes in the female hippocampus conducive to dementia"

### Supplementary Material

#### 1 Supplementary Tables

**Supplementary Table 1. Average age at injury and death of traumatic brain injury (TBI) donors and controls stratified by sex.**

| | Age at TBI | | Age at death* | |
|---|---|---|---|---|
| **Donor category** | **Male** | **Female** | **Male** | **Female** |
| *TBI donors* | | | | |
| Presenting with dementia | 27.63 | 50.00 | 89.27 | 89.26 |
| No dementia | 34.25 | 69.86 | 87.25 | 94.43 |
| *Control donors* | | | | |
| Presenting with dementia | - | - | 89.56 | 92.125 |
| No dementia | - | - | 86.31 | 89.93 |

*Age at death is an approximation, as donors who died between the ages of 90-99 were reported as dying at age "90-94" or "95-99". For individuals in the "90-94" category, an age at death of 92 was used for calculations and an age at death of 97 for the "95-99" category.

Abbreviations: TBI, traumatic brain injury.

**Supplementary Table 2. Genes assigned to the black candidate module from the female hippocampus by weighted co-expression network analysis.**

| Chromosome | Entrez ID | Gene name |
|---|---|---|
| 1 | *ZBTB8B* | zinc finger and BTB domain containing 8B |
| 1 | *ERMAP* | erythroblast membrane-associated protein (Scianna blood group) |
| 1 | *CTBP2P8* | C-terminal binding protein 2 pseudogene 8 |
| 1 | *LOC729987* | uncharacterized LOC729987 |
| 1 | *PMVK* | phosphomevalonate kinase |
| 1 | *DISC2* | disrupted in schizophrenia 2 (non-protein coding) |
| 2 | *LINC01125* | long intergenic non-protein coding RNA 1125 |
| 2 | *CALCRL* | calcitonin receptor-like |
| 3 | *LRTM1* | leucine-rich repeats and transmembrane domains 1 |
| 3 | *ALDH1L1-AS2* | ALDH1L1 antisense RNA 2 |
| 3 | *EPHB1* | EPH receptor B1 |
| 4 | *BANK1* | B-cell scaffold protein with ankyrin repeats 1 |
| 5 | *LOC100287046* | glucuronidase, beta pseudogene |
| 5 | *PSD2* | pleckstrin and Sec7 domain containing 2 |
| 5 | *CCDC69* | coiled-coil domain containing 69 |
| 6 | *LOC105374933* | uncharacterized LOC105374933 |
| 6 | *COL21A1* | collagen, type XXI, alpha 1 |
| 6 | *NT5E* | 5''-nucleotidase, ecto (CD73) |
| 6 | *LOC102723409* | uncharacterized LOC102723409 |

| | | |
|---|---|---|
| 7 | *LOC105375218* | uncharacterized LOC105375218 |
| 7 | *LOC105375515* | uncharacterized LOC105375515 |
| 8 | *REXO1L3P* | REX1, RNA exonuclease 1 homolog (S. cerevisiae)-like 3, pseudogene |
| 9 | *GLDC* | glycine dehydrogenase (decarboxylating) |
| 9 | *PHYHD1* | phytanoyl-CoA dioxygenase domain containing 1 |
| 11 | *HMGN1P21* | high mobility group nucleosome binding domain 1 pseudogene 21 |
| 11 | *TMEM86A* | transmembrane protein 86A |
| 11 | *FAM111B* | family with sequence similarity 111, member B |
| 11 | *LTBP3* | latent transforming growth factor beta binding protein 3 |
| 11 | *PUS3* | pseudouridylate synthase 3 |
| 11 | *KCNJ5* | potassium inwardly-rectifying channel, subfamily J, member 5 |
| 12 | *B3GNT4* | UDP-GlcNAc:betaGal beta-1,3-N-acetylglucosaminyltransferase 4 |
| 12 | *PUS1* | pseudouridylate synthase 1 |
| 13 | *LOC101927248* | uncharacterized LOC101927248 |
| 14 | *SALL2* | spalt-like transcription factor 2 |
| 14 | *IPO4* | importin 4 |
| 14 | *LOC105370549* | uncharacterized LOC105370549 |
| 15 | *C15orf52* | chromosome 15 open reading frame 52 |
| 16 | *LOC105371086* | uncharacterized LOC105371086 |
| 16 | *ORAI3* | ORAI calcium release-activated calcium modulator 3 |
| 16 | *LOC105371305* | uncharacterized LOC105371305 |

| | | |
|---|---|---|
| 16 | *PSKH1* | protein serine kinase H1 |
| 19 | *LOC105372279* | uncharacterized LOC105372279 |
| 19 | *LOC102724908* | uncharacterized LOC102724908 |
| 19 | *NPHS1* | nephrosis 1, congenital, Finnish type (nephrin) |
| 19 | *ZNF749* | zinc finger protein 749 |
| 20 | *RIN2* | Ras and Rab interactor 2 |
| 20 | *ZHX3* | zinc fingers and homeoboxes 3 |
| 21 | *CYYR1-AS1* | cysteine/tyrosine-rich 1 antisense RNA 1 |
| 21 | *SSR4P1* | signal sequence receptor, delta pseudogene 1 |

**Supplementary Table 3. Genes assigned to the green candidate module from the female hippocampus by weighted co-expression network analysis.**

| Chromosome | Entrez ID | Gene name |
|---|---|---|
| 1 | *H6PD* | hexose-6-phosphate dehydrogenase (glucose 1-dehydrogenase) |
| 1 | *LOC105376835* | uncharacterized LOC105376835 |
| 1 | *CCDC163P* | coiled-coil domain containing 163, pseudogene |
| 1 | *CYP4A11* | cytochrome P450, family 4, subfamily A, polypeptide 11 |
| 1 | *GNG5* | guanine nucleotide binding protein (G protein), gamma 5 |
| 1 | *NBPF14* | neuroblastoma breakpoint family, member 14 |
| 1 | *NPL* | N-acetylneuraminate pyruvate lyase (dihydrodipicolinate synthase) |
| 1 | *LOC105371645* | uncharacterized LOC105371645 |
| 2 | *ODC1* | ornithine decarboxylase 1 |
| 2 | *SF3B6* | splicing factor 3b, subunit 6, 14kDa |
| 2 | *OST4* | oligosaccharyltransferase 4 homolog (S. cerevisiae) |
| 2 | *HNRNPA1P39* | heterogeneous nuclear ribonucleoprotein A1 pseudogene 39 |
| 3 | *DZIP1L* | DAZ interacting zinc finger protein 1-like |
| 3 | *SERP1* | stress-associated endoplasmic reticulum protein 1 |
| 3 | *RFC4* | replication factor C (activator 1) 4, 37kDa |
| 3 | *LPP-AS2* | LPP antisense RNA 2 |
| 4 | *UGDH* | UDP-glucose 6-dehydrogenase |
| 4 | *HSD17B13* | hydroxysteroid (17-beta) dehydrogenase 13 |
| 4 | *SLC39A8* | solute carrier family 39 (zinc transporter), member 8 |

| | | |
|---|---|---|
| 4 | *TNRC18P1* | TNRC18P1 |
| 5 | *GUSBP1* | glucuronidase, beta pseudogene 1 |
| 5 | *LOC100132356* | uncharacterized LOC100132356 |
| 5 | *SNCAIP* | synuclein, alpha interacting protein |
| 5 | *CDKL3* | cyclin-dependent kinase-like 3 |
| 5 | *ABLIM3* | actin binding LIM protein family, member 3 |
| 6 | *TUBB2BP1* | tubulin, beta 2B class IIb pseudogene 1 |
| 6 | *LOC105374926* | uncharacterized LOC105374926 |
| 6 | *LOC105374969* | uncharacterized LOC105374969 |
| 6 | *LINC01012* | long intergenic non-protein coding RNA 1012 |
| 6 | *PGM3* | phosphoglucomutase 3 |
| 6 | *NHSL1* | NHS-like 1 |
| 6 | *RPL27AP6* | ribosomal protein L27a pseudogene 6 |
| 6 | *DYNLT1* | dynein, light chain, Tctex-type 1 |
| 6 | *MPC1* | mitochondrial pyruvate carrier 1 |
| 7 | *MACC1* | metastasis associated in colon cancer 1 |
| 7 | *LOC105375222* | uncharacterized LOC105375222 |
| 7 | *STYXL1* | serine/threonine/tyrosine interacting-like 1 |
| 7 | *STAG3L5P-PVRIG2P-PILRB* | STAG3L5P-PVRIG2P-PILRB readthrough |
| 7 | *IQUB* | IQ motif and ubiquitin domain containing |

| | | |
|---|---|---|
| 7 | *LOC105375513* | uncharacterized LOC105375513 |
| 8 | *BTF3P3* | basic transcription factor 3, pseudogene 3 |
| 8 | *ADAM28* | ADAM metallopeptidase domain 28 |
| 8 | *FNTA* | farnesyltransferase, CAAX box, alpha |
| 8 | *LOC100288748* | uncharacterized LOC100288748 |
| 9 | *FREM1* | FRAS1 related extracellular matrix 1 |
| 9 | *LOC730110* | zinc finger protein 492 pseudogene |
| 9 | *MRPS10P5* | mitochondrial ribosomal protein S10 pseudogene 5 |
| 9 | *LOC100499484* | SUGT1-1300002K09Rik pseudogene |
| 9 | *NIPSNAP3A* | nipsnap homolog 3A (C. elegans) |
| 9 | *CTNNAL1* | catenin (cadherin-associated protein), alpha-like 1 |
| 10 | *LOC105376426* | uncharacterized LOC105376426 |
| 10 | *HNRNPA1P32* | heterogeneous nuclear ribonucleoprotein A1 pseudogene 32 |
| 10 | *LOC105378286* | uncharacterized LOC105378286 |
| 10 | *TMEM254-AS1* | TMEM254 antisense RNA 1 |
| 10 | *RNLS* | renalase, FAD-dependent amine oxidase |
| 10 | *BCCIP* | BRCA2 and CDKN1A interacting protein |
| 11 | *KCNQ1* | potassium voltage-gated channel, KQT-like subfamily, member 1 |
| 11 | *TRIM34* | tripartite motif containing 34 |
| 11 | *WDR74* | WD repeat domain 74 |
| 11 | *SLC22A9* | solute carrier family 22 (organic anion transporter), member 9 |

| | | |
|---|---|---|
| 11 | *C11orf65* | chromosome 11 open reading frame 65 |
| 11 | *COLCA1* | colorectal cancer associated 1 |
| 12 | *LOC105369652* | uncharacterized LOC105369652 |
| 12 | *LOC100420899* | mitochondrial ribosomal protein S25 pseudogene |
| 13 | *SLC35E1P1* | solute carrier family 35, member E1 pseudogene 1 |
| 13 | *TEX26* | testis expressed 26 |
| 13 | *CLYBL* | citrate lyase beta like |
| 14 | *SPTSSA* | serine palmitoyltransferase, small subunit A |
| 14 | *LOC105370525* | uncharacterized LOC105370525 |
| 15 | *HERC2P6* | hect domain and RLD 2 pseudogene 6 |
| 15 | *GRAMD2* | GRAM domain containing 2 |
| 15 | *DNM1P9* | DNM1 pseudogene 9 |
| 15 | *VIMP* | VCP-interacting membrane protein |
| 16 | *LOC652276* | potassium channel tetramerization domain containing 5 pseudogene |
| 16 | *NAGPA-AS1* | NAGPA antisense RNA 1 |
| 16 | *ACSM5* | acyl-CoA synthetase medium-chain family member 5 |
| 16 | *HMGN2P3* | high mobility group nucleosomal binding domain 2 pseudogene 3 |
| 16 | *ITGAX* | integrin, alpha X (complement component 3 receptor 4 subunit) |
| 16 | *MYLK3* | myosin light chain kinase 3 |
| 16 | *CBFB* | core-binding factor, beta subunit |

| | | |
|---|---|---|
| 16 | *TXNL4B* | thioredoxin-like 4B |
| 16 | *SNAI3-AS1* | SNAI3 antisense RNA 1 |
| 17 | *ANKFN1* | ankyrin-repeat and fibronectin type III domain containing 1 |
| 17 | *LINC01152* | long intergenic non-protein coding RNA 1152 |
| 19 | *LOC105372293* | uncharacterized LOC105372293 |
| 19 | *RPS19* | ribosomal protein S19 |
| 19 | *ZNF222* | zinc finger protein 222 |
| 20 | *TGIF2* | TGFB-induced factor homeobox 2 |
| 22 | *SLC35E4* | solute carrier family 35, member E4 |
| X | *MTHFD1P1* | methylenetetrahydrofolate dehydrogenase (NADP+ dependent) 1 pseudogene 1 |

**Supplementary Table 4. Genes assigned to the red candidate module from the female hippocampus by weighted co-expression network analysis.**

| Chromosome | Entrez ID | Gene name |
|---|---|---|
| 1 | *PINK1-AS* | PINK1 antisense RNA |
| 1 | *CD84* | CD84 molecule |
| 1 | *LOC105371602* | uncharacterized LOC105371602 |
| 1 | *SLC30A1* | solute carrier family 30 (zinc transporter), member 1 |
| 1 | *LOC101927765* | uncharacterized LOC101927765 |
| 1 | *PSMD2P1* | proteasome 26S subunit, non-ATPase, 2 pseudogene 1 |
| 2 | *ZFP36L2* | ZFP36 ring finger protein-like 2 |
| 2 | *LOXL3* | lysyl oxidase-like 3 |
| 2 | *YWHAQP5* | YWHAQ pseudogene 5 |
| 2 | *MERTK* | MER proto-oncogene, tyrosine kinase |
| 2 | *PTPN18* | protein tyrosine phosphatase, non-receptor type 18 (brain-derived) |
| 3 | *TMEM43* | transmembrane protein 43 |
| 3 | *LOC105376974* | uncharacterized LOC105376974 |
| 4 | *LOC105377299* | uncharacterized LOC105377299 |
| 4 | *LOC105377319* | uncharacterized LOC105377319 |
| 5 | *FAM105A* | family with sequence similarity 105, member A |
| 5 | *ADRB2* | adrenoceptor beta 2, surface |
| 6 | *RUNX2* | runt-related transcription factor 2 |

| | | |
|---|---|---|
| 6 | *LINC01268* | long intergenic non-protein coding RNA 1268 |
| 6 | *LOC100129577* | mitochondrial carrier 1 pseudogene |
| 7 | *LOC100506178* | uncharacterized LOC100506178 |
| 7 | *AOAH* | acyloxyacyl hydrolase (neutrophil) |
| 7 | *LOC105375532* | uncharacterized LOC105375532 |
| 7 | *LOC105375531* | uncharacterized LOC105375531 |
| 9 | *LOC101927069* | uncharacterized LOC101927069 |
| 9 | *SYK* | spleen tyrosine kinase |
| 10 | *LOC105376427* | uncharacterized LOC105376427 |
| 10 | *LOC105376425* | uncharacterized LOC105376425 |
| 10 | *TRDMT1* | tRNA aspartic acid methyltransferase 1 |
| 10 | *LRRC37A6P* | leucine rich repeat containing 37, member A6, pseudogene |
| 10 | *LOC105378280* | uncharacterized LOC105378280 |
| 10 | *LOC105378294* | uncharacterized LOC105378294 |
| 10 | *LOC105378367* | uncharacterized LOC105378367 |
| 10 | *HOGA1* | 4-hydroxy-2-oxoglutarate aldolase 1 |
| 10 | *WDR11-AS1* | WDR11 antisense RNA 1 |
| 12 | *CLEC7A* | C-type lectin domain family 7, member A |
| 12 | *PCED1B-AS1* | PCED1B antisense RNA 1 |
| 12 | *FMNL3* | formin-like 3 |
| 14 | *PSME1* | proteasome (prosome, macropain) activator subunit 1 (PA28 alpha) |

| | | |
|---|---|---|
| 14 | *C14orf105* | chromosome 14 open reading frame 105 |
| 15 | *LRRK1* | leucine-rich repeat kinase 1 |
| 16 | *CIITA* | class II, major histocompatibility complex, transactivator |
| 16 | *LOC105371240* | uncharacterized LOC105371240 |
| 16 | *MMP2* | matrix metallopeptidase 2 (gelatinase A, 72kDa gelatinase, 72kDa type IV collagenase) |
| 17 | *PLXDC1* | plexin domain containing 1 |
| 17 | *TMEM106A* | transmembrane protein 106A |
| 17 | *SLC16A6P1* | SLC16A6 pseudogene 1 |
| 18 | *RPL10AP13* | ribosomal protein L10a pseudogene 13 |
| 19 | *ZNF846* | zinc finger protein 846 |
| 21 | *B3GALT5* | UDP-Gal:betaGlcNAc beta 1,3-galactosyltransferase, polypeptide 5 |
| X | *CSF2RA* | colony stimulating factor 2 receptor, alpha, low-affinity (granulocyte-macrophage) |

**Supplementary Table 5. P-values from the comparison of expression levels of genes of interest between traumatic brain injury (TBI) donors presenting with dementia and all other categories of donors, stratified by sex.**

| | **P-value** | | | | | |
|---|---|---|---|---|---|---|
| | **Female: TBI dementia vs.** | | | **Male: TBI dementia vs.** | | |
| **Gene of interest** | **TBI no dementia** | **Control no dementia** | **Control dementia** | **TBI no dementia** | **Control no dementia** | **Control dementia** |
| *ACSM5* | 0.064 | 0.016 | 0.0092 | 0.49 | 0.60 | 0.47 |
| *ADAM28* | 0.0015 | 0.00066 | 0.0047 | 0.45 | 0.91 | 0.84 |
| *CCDC163P* | 0.0033 | 0.02 | 0.00052 | 0.043 | 0.24 | 0.56 |
| *COLCA1* | 0.0045 | 0.44 | 0.17 | 0.33 | 0.44 | 0.28 |
| *FMNL3* | 0.012 | 0.03 | 0.18 | 0.93 | 0.68 | 0.90 |
| *LOC105371645* | 0.019 | 0.21 | 0.13 | 0.36 | 0.35 | 0.90 |
| *NPL* | 0.033 | 0.02 | 0.24 | 0.76 | 0.29 | 0.51 |
| *ORAI3* | 0.022 | 0.037 | 0.24 | 0.28 | 0.073 | 0.47 |
| *RIN2* | 0.019 | 0.00094 | 0.087 | 0.56 | 0.14 | 0.65 |
| *ANKFN1* | 0.012 | 0.03 | 0.029 | 0.80 | 0.56 | 0.51 |
| *PTPN18* | 0.019 | 0.013 | 0.072 | 0.30 | 0.95 | 0.79 |
| *ZHX3* | 0.00066 | 0.16 | 0.017 | 0.095 | 0.064 | 0.47 |

Statistical analyses were conducted using Wilcoxon's Rank Sum test. Significance was measured at an alpha-level of 0.05.

**Supplementary Table 6. Gene ontology biological process annotated to the black candidate module from the female hippocampus.**

Please see Supplementary Table 6 Excel file.

**Supplementary Table 7. Gene ontology biological process annotated to the green candidate module from the female hippocampus.**

Please see Supplementary Table 7 Excel file.

**Supplementary Table 8. Gene ontology biological process annotated to the red candidate module from the female hippocampus.**

Please see Supplementary Table 8 Excel file.

### 2 Supplementary Figures

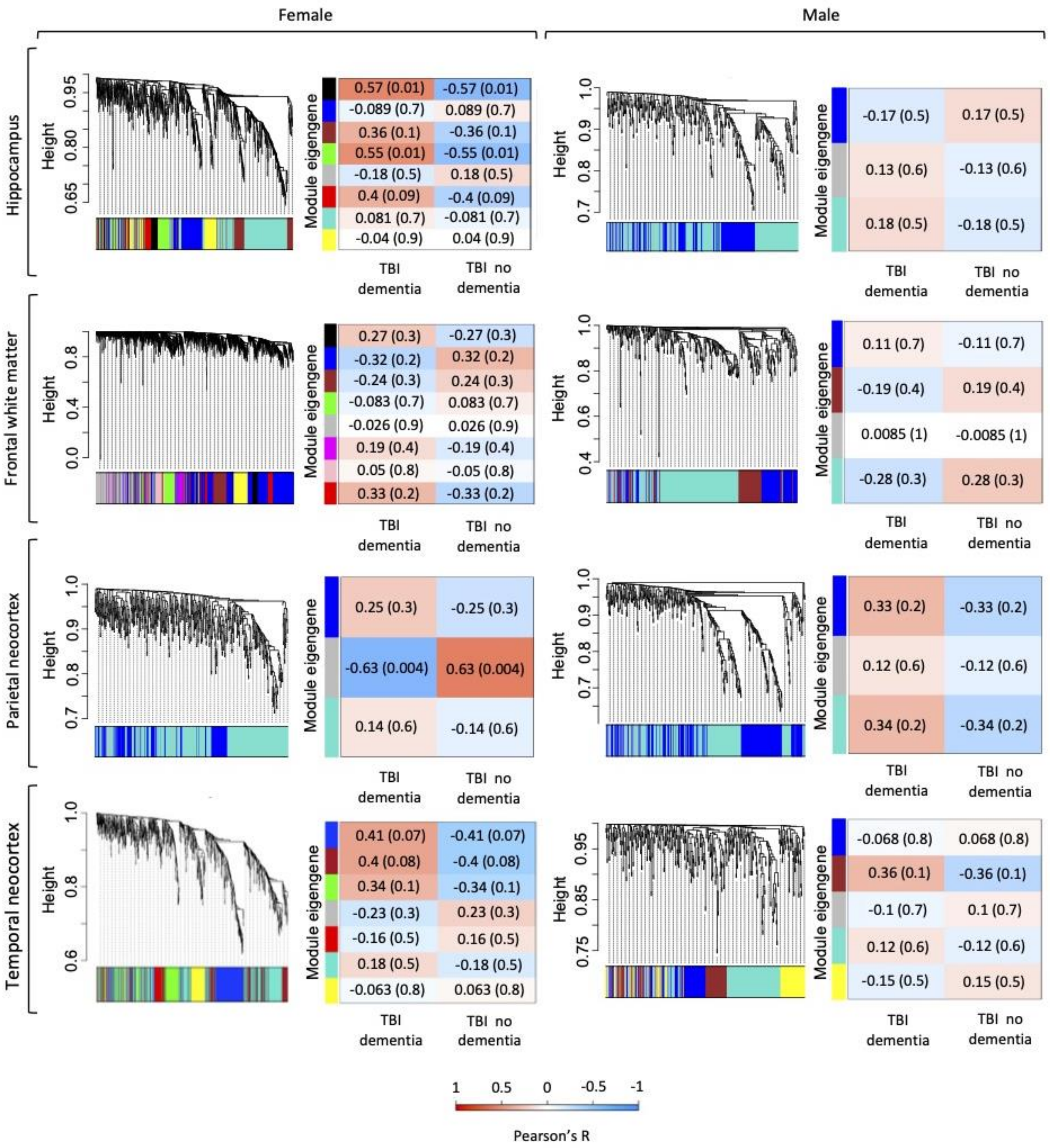


**Supplementary Figure 1. Summary of weighted gene co-expression network analyses of TBI-associated differentially expressed genes stratified by sample sex and brain region source.** For each analysis, the co-expression cluster dendrogram of TBI-associated differentially expressed genes (DEGs) (left) and the module-trait relationship with dementia status in TBI donors (right) are shown. Correlation coefficients and p-values (in brackets) are indicated in each respective cell. Abbreviations: TBI, traumatic brain injury.

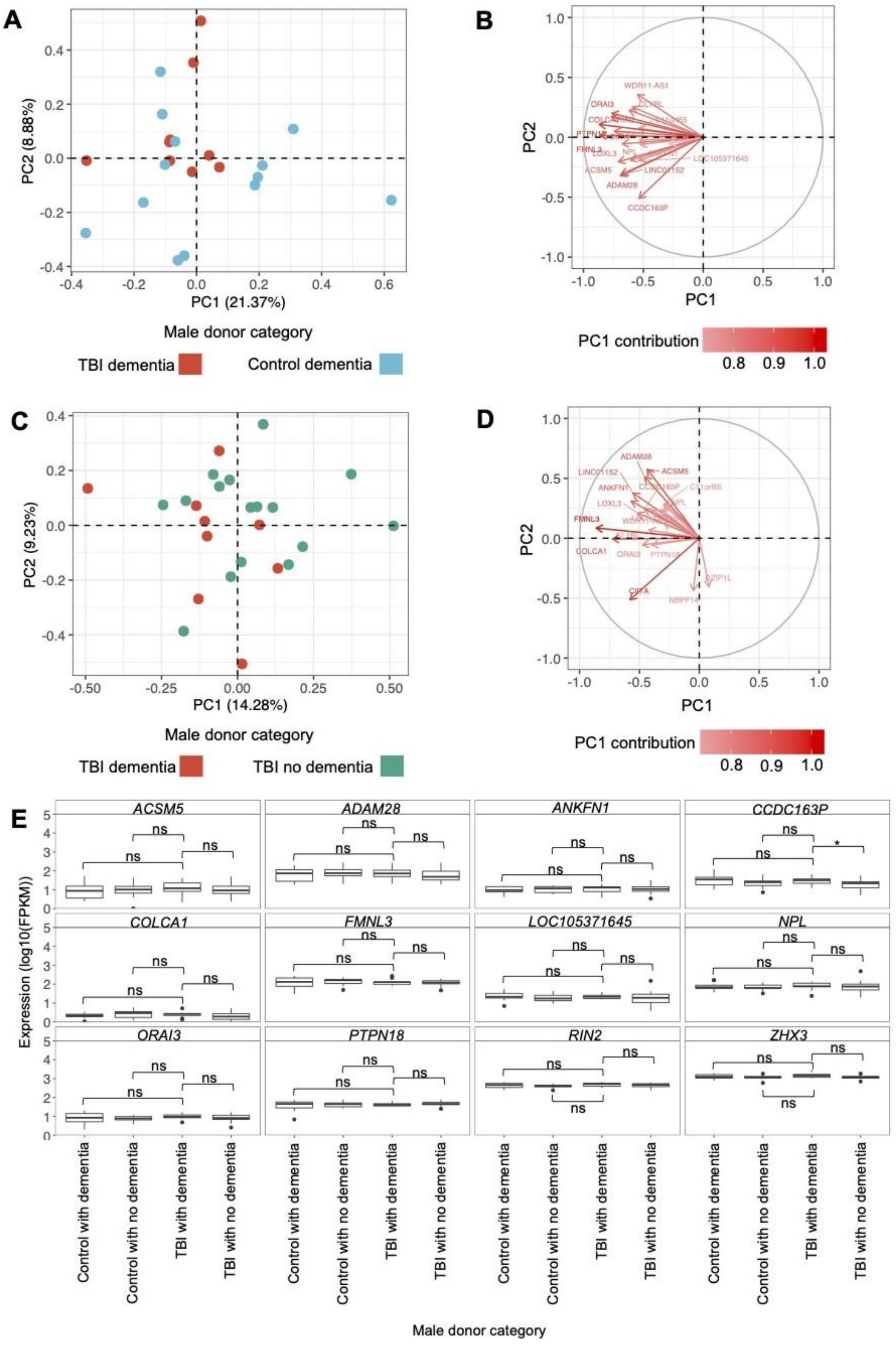
A
PC1 (21.37%)
PC2 (8.88%)
Male donor category
TBI dementia
Control dementia
B
PC1
PC2
PC1 contribution
C
PC1 (14.28%)
PC2 (9.23%)
Male donor category
TBI dementia
TBI no dementia
D
PC1
PC2
PC1 contribution
E
ACSM5
ADAM28
ANKFN1
CCDC163P
COLCA1
FMNL3
LOC105371645
NPL
ORAI3
PTPN18
RIN2
ZHX3
Expression (log10(FPKM))
Control with dementia
Control with no dementia
TBI with dementia
TBI with no dementia
Male donor category

**Supplementary Figure 2. Principal component analysis (PCA) and gene-protein correlations in the male hippocampus of candidate modules identified from the female hippocampus. (A)** PCA using fragments per kilobase of transcript per million mapped reads (FPKM) of the genes assigned to candidate modules from the female hippocampus, in the hippocampus of male TBI donors presenting with dementia and male controls presenting with dementia. The primary and secondary PCs are shown, and their contribution to the variation is indicated on the respective axis. **(B)** Eigenvectors and percent contribution of the top 20 genes showing the highest contribution to PC1 in the female hippocampus PCA1, which compared the expression of candidate module genes in the female hippocampus between TBI donors presenting with dementia and controls presenting with dementia. Here, the percent contribution and eigenvectors of these genes are displayed for their contribution to the male PCA shown in plot A. Progressively darker red color indicates higher contribution to PC1. **(C)** PCA using FPKM of the genes assigned to candidate modules from the female hippocampus, in the hippocampus of male TBI donors presenting with dementia and male TBI donors not presenting with dementia. **(D)** Eigenvectors and percent contribution of the top 20 genes showing the highest contribution to PC1 in the female hippocampus PCA2, which compared the expression of candidate module genes in the female hippocampus between TBI donors presenting with dementia and TBI donors not presenting with dementia. Here, the percent contribution and eigenvectors of these genes are displayed for their contribution to the male PCA shown in plot C. **(E)** Expression levels of genes of interest— including primary hub genes (*ANKFN1*, *PRPN18*, *ZHX3*) and genes that were amongst the top 20 contributors to PC1 in both female hippocampus PCAs — in the male hippocampus were compared using the Wilcoxon Rank Sum test between male TBI donors presenting with dementia and all other categories of male donors. Within each panel, boxplots for each category of male donor are displayed in the same order as the figure legend. * $p < 0.05$, ** $p < 0.01$, *** $p < 0.001$, ns = not significant. Abbreviations: FPKM, fragments per kilobase of transcript per million mapped reads; PC1; primary principal component; PC2, secondary principal component; TBI, traumatic brain injury.
